# Risk factors associated with development and persistence of long COVID

**DOI:** 10.1101/2021.09.22.21263998

**Authors:** Yusuke Miyazato, Shinya Tsuzuki, Shinichiro Morioka, Mari Terda, Satoshi Kutsuna, Sho Saito, Yumiko Shimanishi, Kozue Takahashi, Mio Sanada, Masako Akashi, Chika Kuge, Yasuyo Osanai, Keiko Tanaka, Michiyo Suzuki, Kayoko Hayakawa, Norio Ohmagari

**Author notes:** Corresponding author: Shinichiro Morioka, Address: 1-21-1 Toyama, Shinjuku-ku, Tokyo 162-8655, Japan. Alternate corresponding author: Yusuke Miyazato, Address: 1-21-1 Toyama, Shinjuku-ku, Tokyo 162-8655, Japan. **Author contributions:** Yusuke Miyazato: Conceptualization, Methodology, Formal analysis, Investigation, Data Curation, Writing-original draft preparation. Shinya Tsuzuki: Conceptualization, Methodology, Formal analysis, Data Curation. Shinichiro Morioka: Conceptualization, Methodology, Formal analysis, Investigation, Data Curation, Writing-original draft preparation, Writing review and editing, Visualization, Supervision, Project administration, Funding acquition. Mari Terda: Conceptualization, Data Curation, Writing-original draft preparation. Satoshi Kutsuna: Conceptualization, Methodology. Sho Saito: Conceptualization, Methodology. Yumiko Shimanishi: Investigation, Data Curation. Kozue Takahashi: Investigation, Data Curation. Mio Sanada: Investigation, Data Curation. Masako Akashi: Conceptualization. Chika Kuge: Conceptualization. Yasuyo Osanai: Conceptualization. Keiko Tanaka: Conceptualization. Michiyo Suzuki: Conceptualization. Kayoko Hayakawa: Conceptualization, Methodology. Norio Ohmagari: Conceptualization, Supervision.

## Abstract

**Background:** Long COVID has been a social concern. Though patient characteristics associated with developing long COVID are partially known, those associated with persisting it have not been identified.

**Methods:** We conducted a cross-sectional questionnaire survey of patients after COVID-19 recovery who visited the National Center for Global Health and Medicine between February 2020 and March 2021. Demographic and clinical data, and the presence and duration of long COVID were obtained. We identified factors associated with development and persistence of long COVID using multivariate logistic and linear regression analysis, respectively.

**Results:** We analyzed 457 of 526 responses (response rate, 86.9%). The median age was 47 years, and 378 patients (84.4%) had mild disease in acute phase. The number of patients with any symptoms after 6 and 12 months after onset or diagnosis were 120 (26.3%) and 40 (8.8%), respectively. Women were at risk for development of fatigue (OR 2.03, 95% CI 1.31-3.14), dysosmia (OR 1.91, 95% CI 1.24-2.93), dysgeusia (OR 1.56, 95% CI 1.02-2.39), and hair loss (OR 3.00, 95% CI 1.77-5.09), and were at risk for persistence of any symptoms (coefficient 38.0, 95% CI 13.3-62.8). Younger age and low body mass index were risk factors for developing dysosmia (OR 0.96, 95% CI 0.94-0.98, and OR 0.94, 95% CI 0.89-0.99, respectively) and dysgeusia (OR 0.98, 95% CI 0.96-1.00, and OR 0.93, 95% CI 0.88-0.98, respectively).

**Conclusion:** We identified risk factors for the persistence as well as development of long COVID. Many patients suffer from long-term residual symptoms, even in mild cases.

**Summary:** Our cross-sectional questionnaire survey of patients recovering from COVID-19 revealed that women, young age, and low body mass index were risk factors for the development of multiple symptoms, and that even mild cases of COVID-19 suffered from long-term residual symptoms.

## Introduction

COVID-19 has become a global threat, with 226 million infections and 4.65 million deaths worldwide as of September 16, 2021 [1]. After the onset of COVID-19, approximately 76% of patients are known to have prolonged symptoms lasting more than six months [2] known as long COVID or post-acute sequelae of SARS-CoV-2 infection (PASC). Previous studies have shown that frequent symptoms of long COVID include fatigue, dyspnea, and dysosmia, and that multiple symptoms often overlap and persist [2] [3] [4]. Some symptoms of COVID-19, such as hair loss, which were not present at the time of onset, may appear after recovery [4]. In addition, it has been reported that 5-10% of patients continue to experience moderate to severe problems at work, and in social and family life, eight months after the onset of COVID-19 [5]. The impact of long COVID on society is immeasurable. However, the characteristics of patients who are more likely to develop long COVID are only partially known [2], and it is important to identify risk factors in order to properly understand and prevent long COVID. In this study, we explored the risk factors for the development and persistence of long COVID in a cohort of patients recovering from COVID-19 at a hospital designated for infectious diseases in Japan.

## Methods

This study was designed as a single-center, cross-sectional survey, in which a self-report questionnaire was mailed to eligible patients in April 2021 with two reminders: two weeks and one month later. Participation in this survey was voluntary, but not anonymous. The participants after COVID-19 recovery were requested to complete and return the questionnaire. Informed consent was confirmed by marking the consent checkbox on the questionnaire. This study was reviewed and approved by the Ethics Committee of the Center Hospital of the National Center for Global Health and Medicine (NCGM) (NCGM-G-004121-00).

### Participants

Potential participants were recruited from patients who had recovered from COVID-19 and visited the outpatient service of the Disease Control and Prevention Center (DCC) in the NCGM from February 2020 to March 2021 in order to obtain a pre-donation screening test for COVID-19 convalescent plasmapheresis [6].

### Questionnaire

We developed the questionnaire through a systematic literature review with reference to similar previous studies [2] [3] [7] [8] [9] [10] [11], findings from our previous study on prolonged and late-onset symptoms of COVID-19 [4], and comprehensive discussions among the authors. We attempted to minimize the number of questions to maximize the response rate. The instrument was piloted on 13 patients who had recovered from COVID-19; they provided feedback on the content, clarity, and format of the items. They also confirmed that the survey questions were self-explanatory. Minor revisions were made in response to their feedback.

### Items investigated

Patient characteristics (e.g., age, sex, height, weight, smoking history, drinking history, underlying medical conditions, and obstetric history), information regarding the acute phase of COVID-19 (e.g., presence of pneumonia, disease severity; oxygen therapy/mechanical ventilation*/*extracorporeal membrane oxygenation: ECMO, treatment, antivirals/steroids), and presence and duration of symptoms related to COVID-19 were investigated (**Appendix 1**). Underlying medical conditions included hypertension, diabetes, dyslipidemia, bronchial asthma, chronic obstructive pulmonary disease (COPD), myocardial infarction, malignancies, connective tissue/rheumatic diseases, immune-deficiency disease, chronic kidney disease, and others. Disease severity was categorized as follows [2] [12]: 1) Mild: no oxygen therapy; 2) Moderate: oxygen therapy without mechanical ventilation; and 3) Severe: mechanical ventilation with or without ECMO. Symptoms related to COVID-19 were fever, fatigue, shortness of breath (SoB), joint pain, myalgia, chest pain, cough, abdominal pain, dysosmia, dysgeusia, runny nose, conjunctivitis, headache, sputum, sore throat, diarrhea, nausea, loss of appetite, hair loss, depression, loss of concentration, and memory disturbance (MD).

### Statistical analysis

The patient characteristics, presence of pneumonia, disease severity, and treatment in the acute phase of COVID-19 were expressed as median, interquartile range (IQR), or % (n), where applicable. The proportion of patients with prolonged symptoms and days since the onset of COVID-19 was described. The symptoms of more than 100 participants in this study were categorized and defined as follows: 1) Acute symptoms, which resolved or did not appear within 28 days after the onset of COVID-19 in >90% of participants; 2) On-going/chronic symptoms, which did not resolve within 28 days after the onset of COVID-19 in >10% of participants; and3) Late-onset symptoms, which appeared 28 days after the onset of COVID-19 in >5% of participants.

Multivariate logistic regression analyses were performed to calculate the adjusted odds ratios (ORs) with 95% confidence intervals (CIs) for the development of on-going/chronic symptoms and late-onset symptoms. The multivariate logistic regression model was performed with adjustments for all potential confounding factors of the participants’ characteristics, presence of pneumonia, disease severity, and treatment in the acute phase of COVID-19. Data were analyzed using Stata BE 17.0 (StataCorp, College Station, TX, USA) and R, version 4.0.5 (R Foundation for Statistical Computing; 2018, Vienna, Austria).

Then, linear regression statistical tests were performed to identify factors associated with the persistence of each symptom among patients who developed the symptom, and the persistence of any symptom associated with COVID-19. The level of significance for all statistical tests was set at α=0.05. The dependent variable was the duration of each symptom (day). We included the participants’ characteristics, presence of pneumonia, disease severity, and treatment in the acute phase of COVID-19 as independent variables. Data were analyzed using Stata BE 17.0 (StataCorp, College Station, TX, USA) and R, version 4.0.5 (R Foundation for Statistical Computing; 2018, Vienna, Austria).

## Results

A self-reported questionnaire was mailed to 526 patients who had recovered from COVID-19, and we obtained 457 responses (response rate, 86.9%). The demographic and clinical characteristics of the participants are summarized in **Table 1**. The median age was 47 years, and 49.5% of the patients were women. All the participants were Japanese. A total of 245 patients (53.7%) had no underlying medical conditions. A total of 173 patients (40.5%) had pneumonia. Regarding severity, 378 patients (84.4%) were mild, 73 (12.7%) were moderate, and 13 (2.9%) were severe. The median number of days from symptom onset or diagnosis of COVID-19 to interview was 248.5 days.

**Table 1.**
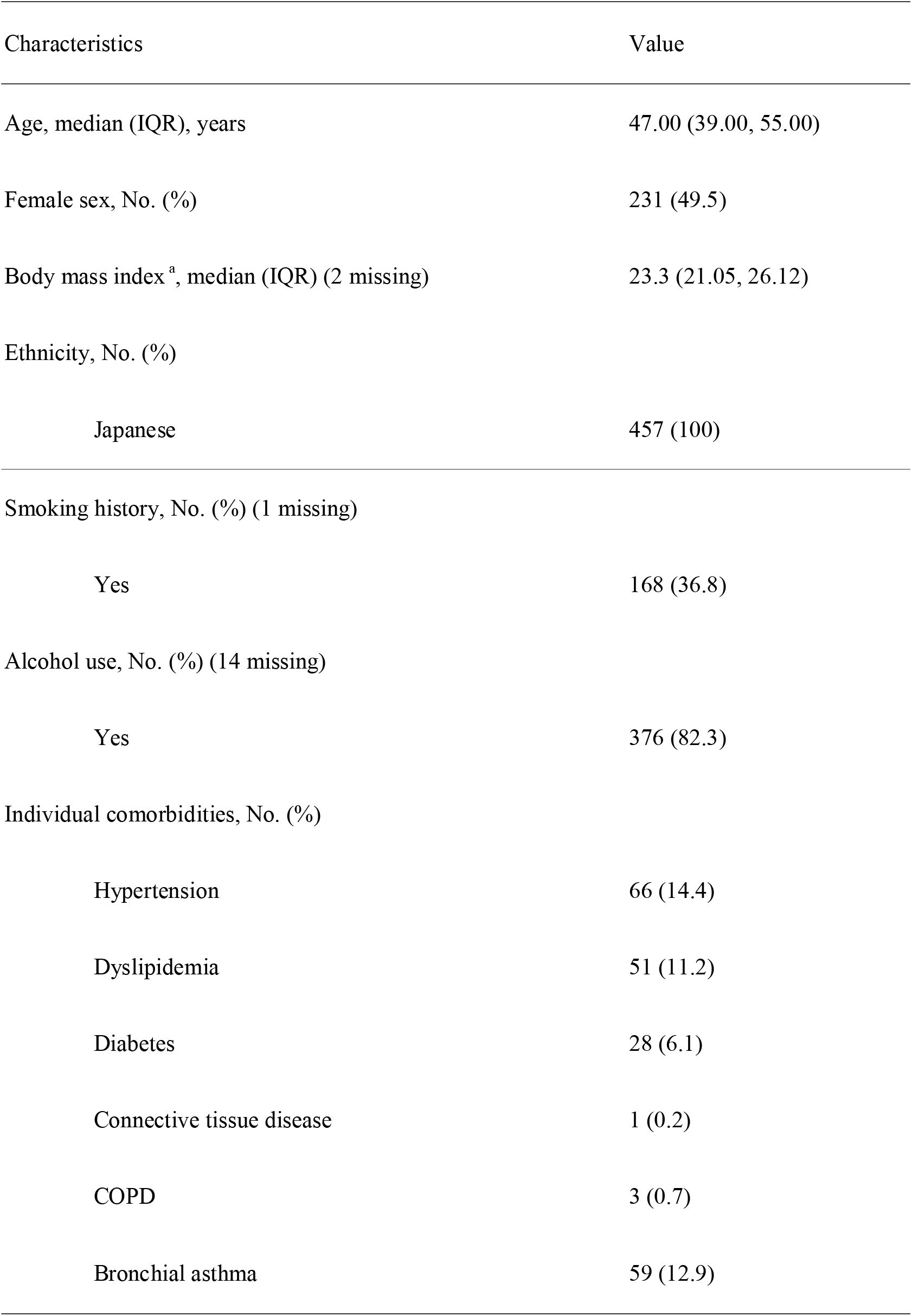
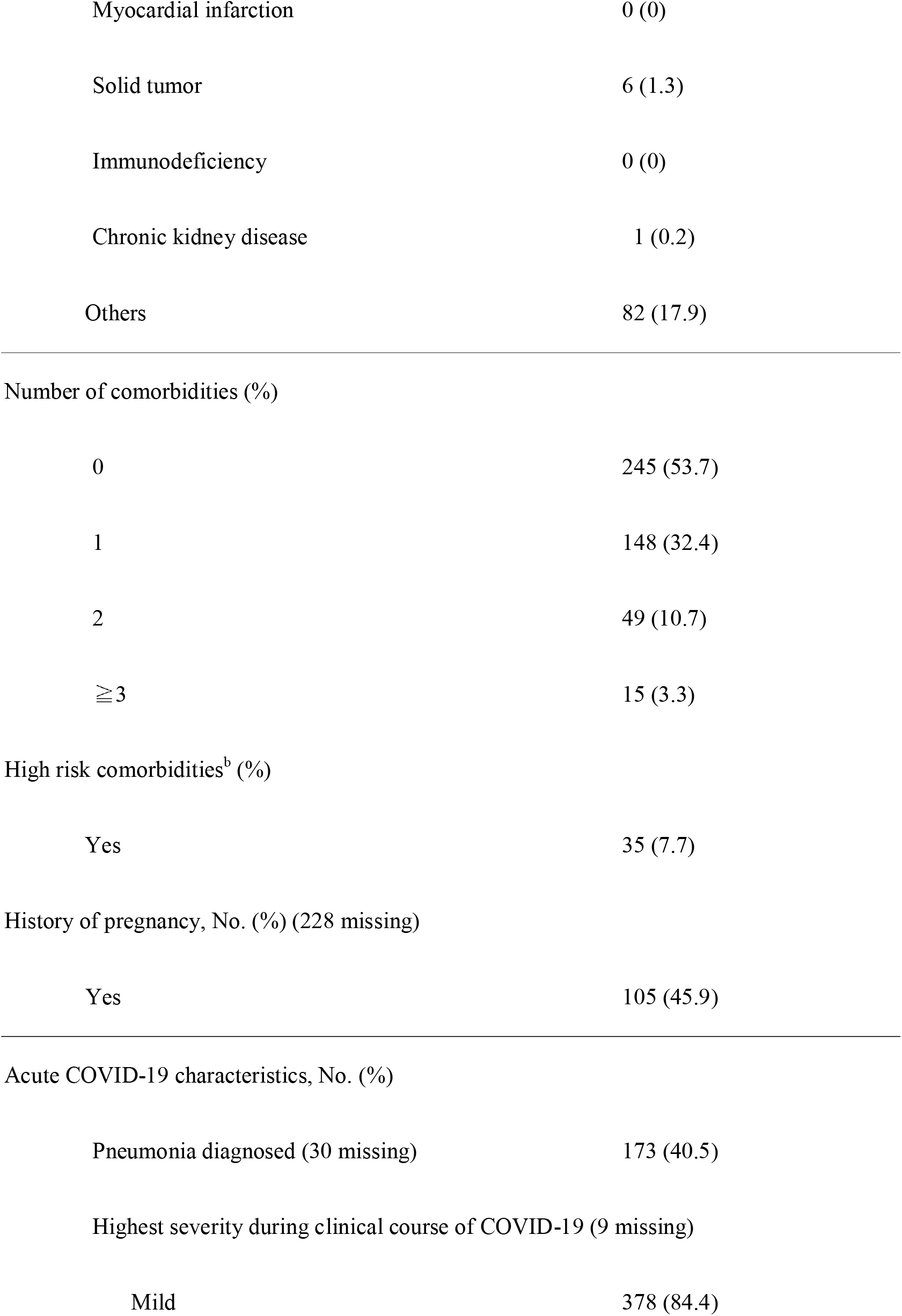
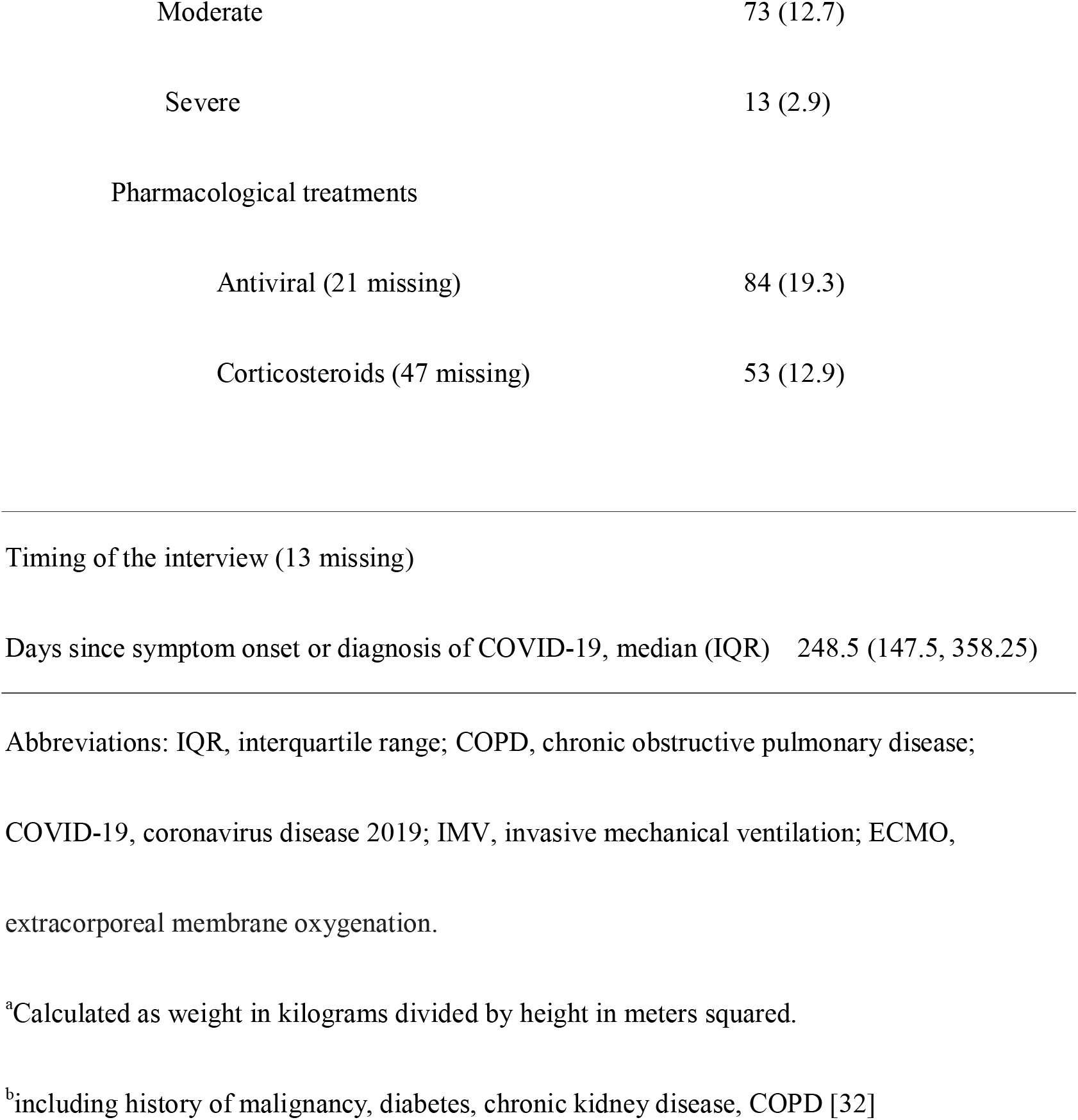
Demographic and clinical characteristics of the participants (n=457)

### Classification of symptoms associated with COVID-19

The number of participants experiencing symptoms associated with COVID-19, the number of participants with symptoms lasting for more than four weeks and 12 weeks, and the number of the participants whose symptoms developed more than four weeks after the onset of COVID-19 are summarized in **Appendix 2**. The most common symptom associated with acute COVID-19 was fever (293 participants, 64.1%), followed by fatigue (292 participants, 64.0%),dysosmia (219 participants, 47.9%), cough (214 participants, 46.8%), and dysgeusia (185 participants, 40.6%). On the other hand, the most common symptom lasting for more than four weeks was dysosmia (104 participants, 22.8%), followed by loss of concentration (95 participants, 20.8%), and fatigue (93 participants, 20.4%). The most common symptom that developed more than four weeks after the onset of COVID-19 was hair loss (58 participants), followed by memory disturbance (17 participants), and depression (16 participants).

We classified 17 long COVID symptoms into three categories after excluding less frequent symptoms reported in less than 100 participants (chest pain, abdominal pain, runny nose, conjunctivitis, and nausea): 1) acute symptoms (e.g., fever, headache, loss of appetite, joint pain, sore throat, myalgia, diarrhea, and sputum) that persisted for four weeks in less than 10% of the participants; 2) On-going/Chronic symptoms (e.g., fatigue, dysosmia, cough, dysgeusia, shortness of breath) that persisted for four weeks in more than 10% of the participants except for late-onset symptoms; and 3) late-onset symptoms (e.g., loss of concentration, depression, hair loss, and memory disturbance), which developed more than four weeks after the onset of COVID-19 in more than 5% of the participants with the symptoms.

The frequency and duration of acute symptoms, on-going/chronic symptoms, and late-onset symptoms are summarized in **Appendix 3, Figure 1** and **2**, respectively. A few participants reported on-going/chronic symptoms, including dysosmia (n=35, 7.7%), fatigue (n=30, 6.6%), shortness of breath (n=18, 3.9%), dysgeusia (n=16, 3.5%), and cough (n=11, 2.4%) six months after symptom onset or diagnosis of COVID-19, and fatigue (n=14, 3.1%), shortness of breath (n=7, 1.5%), dysosmia (n=5, 1.1%), cough (n=5, 1.1%), and dysgeusia (n=2, 0.4%) 12 months after symptom onset or diagnosis of COVID-19. The participant with the longest duration of ongoing/chronic symptoms was a 43-year-old woman who had been suffering from shortness of breath, cough, and fatigue for 439 days. Late-onset symptoms were also persistent: memory disturbance (n=52, 11.4%), loss of concentration (n=45, 9.8%), depression (n=37, 8.1%), and hair loss (n=14, 3.1%) six months after symptom onset or diagnosis of COVID-19, and memory disturbance (n=25, 5.5%), loss of concentration (n=22, 4.8%), depression (n=15, 3.3%), and hair loss (n=2, 0.4%) 12 months after symptom onset or diagnosis of COVID-19. Among the 89 patients who developed hair loss (14 missing), 83 participants (93.3%) had diffuse hair loss, and 6 participants (6.7%) had patchy hair loss.

**Figure 1.**
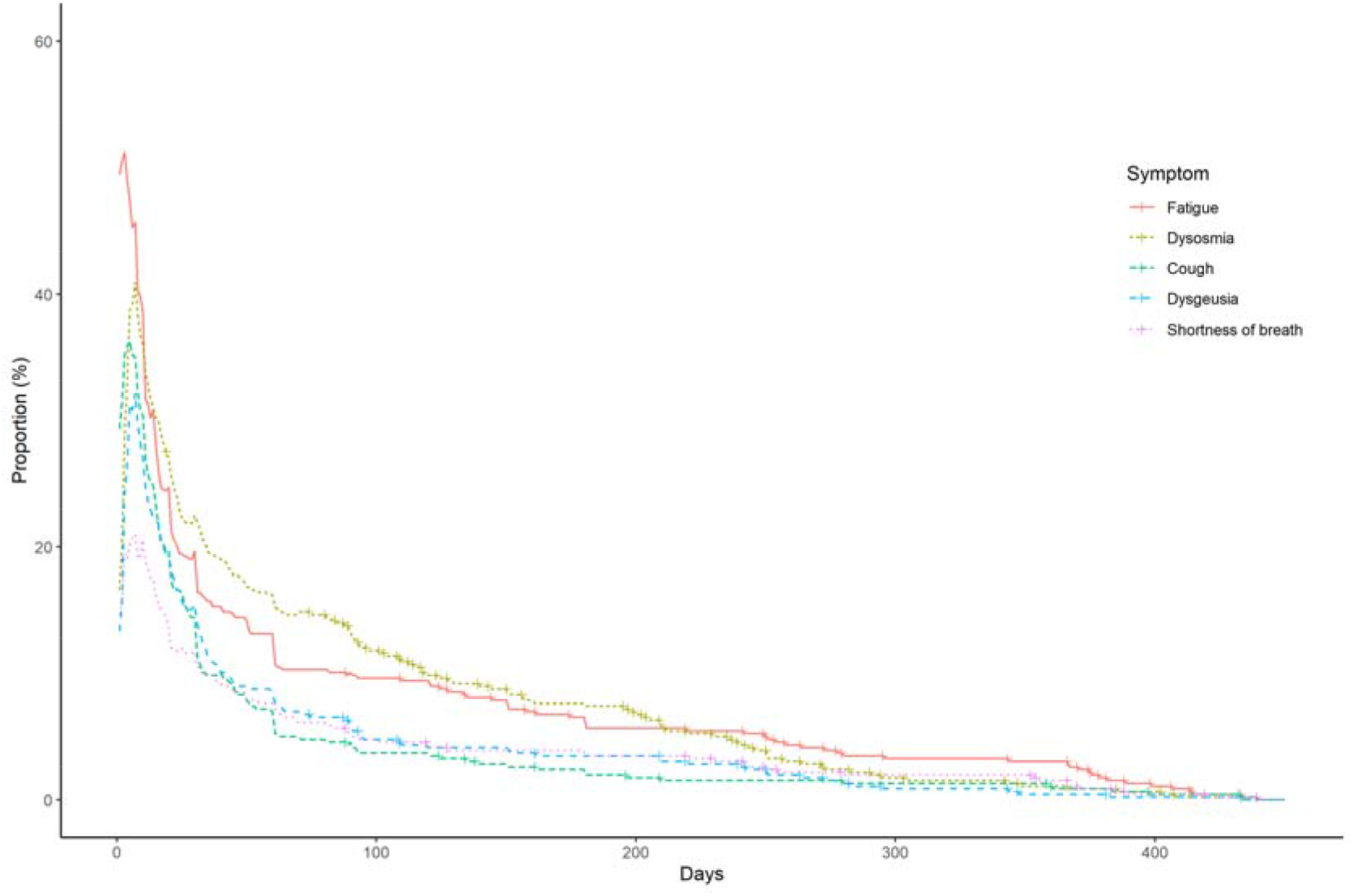
Ongoing/chronic symptoms associated with COVID-19.

**Figure 2.**
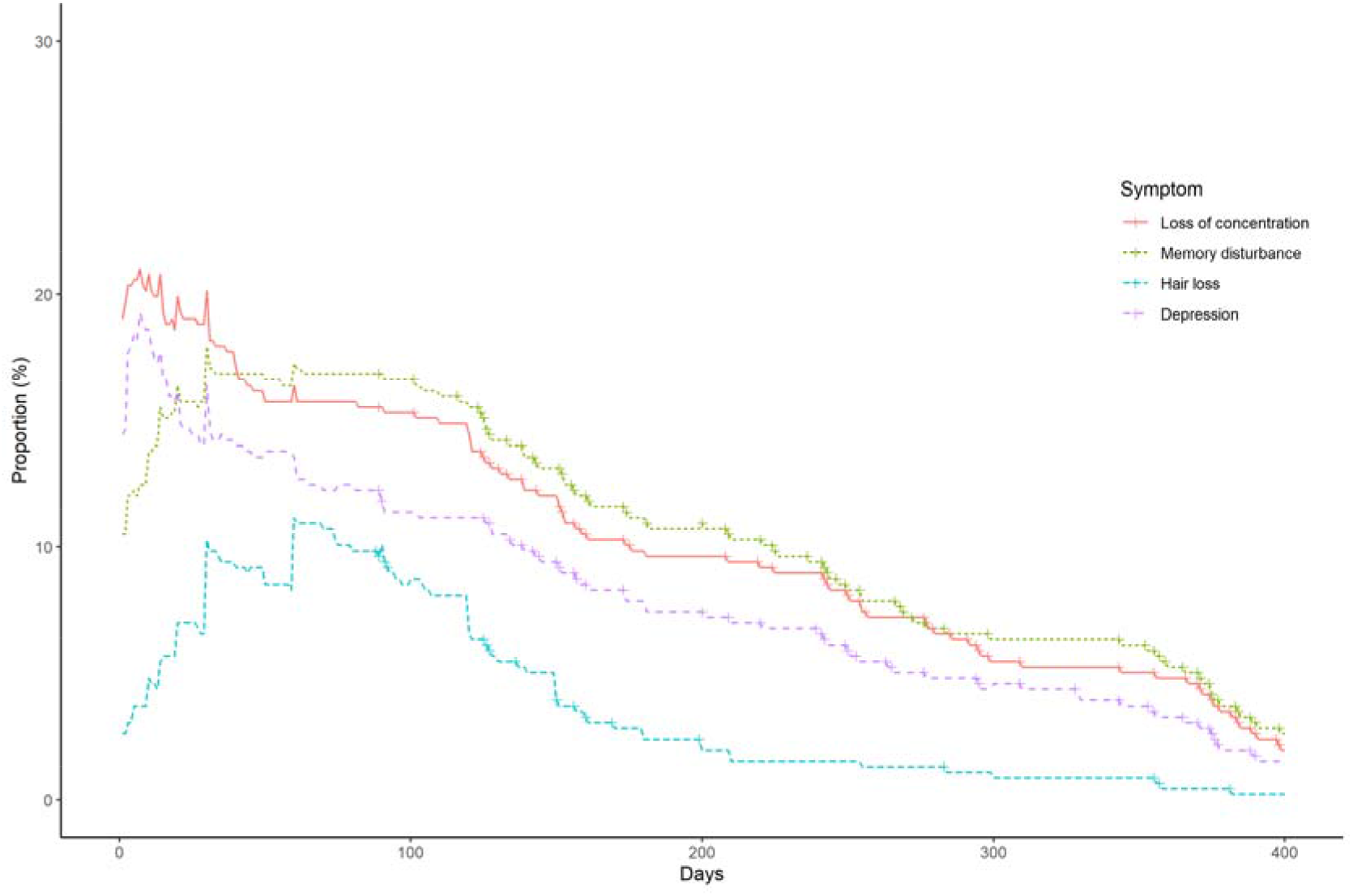
Late-onset symptoms associated with COVID-19.

Moreover, the frequency and duration of at least one symptom are shown in **Figure 3** (any symptoms included acute, on-going/chronic, and late-onset symptoms). The number of participants with at least one symptom after six months and 12 months after symptom onset or diagnosis of COVID-19 were 120 (26.3%) and 40 (8.8%), respectively.

**Figure 3.**
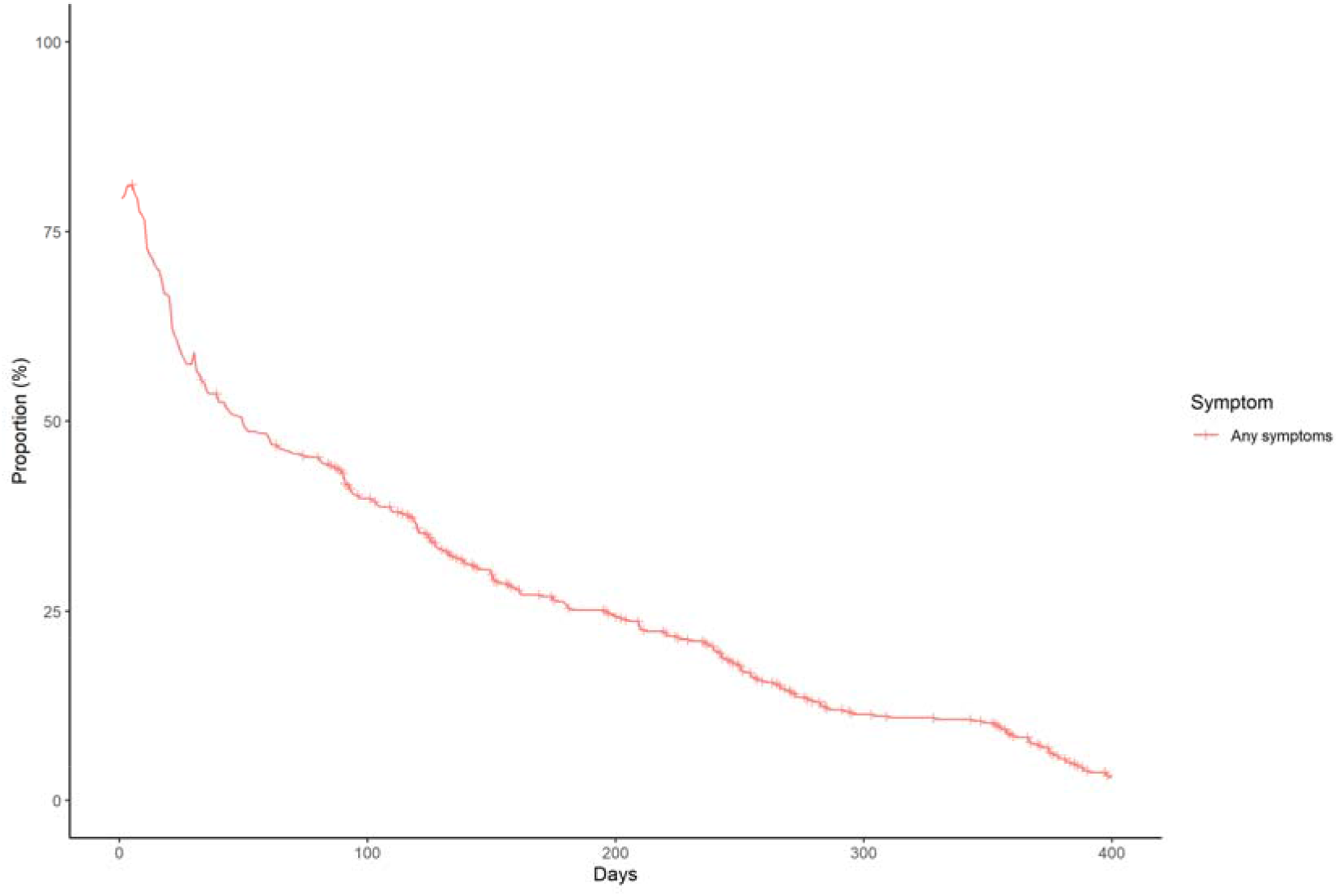
The proportion of the participants who had at least one symptom associated with COVID-19.

We identified the risk factors for the development and persistence of some of the common symptoms: fatigue, dysgeusia, dysosmia, and hair loss. After multivariable adjustment, development of fatigue was associated with female sex (OR 2.03, *p*=0.001, 95% CI 1.31-3.14), while persistence of fatigue among patients with fatigue was associated with being diagnosed with pneumonia (coefficient 17.3, *p*=0.028, 95% CI 1.92-32.6), and moderate severity compared to mild severity (coefficient 41.5, *p*=0.033, 95% CI 3.33-79.7) (**Appendix 4a, 4b**). The risk of developing dysgeusia was associated with female sex (OR 1.56, *p*=0.042, 95% CI 1.02-2.39), and was inversely associated with age (OR 0.98, *p*=0.015, 95% CI 0.96-1.00) and BMI (OR 0.93, *p*=0.012, 95% CI 0.88-0.98). Persistence of dysgeusia among patients with dysgeusia was associated with female sex (coefficient 28.7, *p*=0.038, 95% CI 1.65-55.7) (**Appendix 5a, 5b**). The risk of developing dysosmia was also associated with female sex (OR 1.91, *p*=0.003, 95% CI 1.24-2.93), and was inversely associated with age (OR 0.96, *p*<0.001, 95% CI 0.94-0.98), BMI (OR 0.94, *p*=0.014, 95% CI 0.89-0.99), and antiviral drug use (OR 0.59, *p*=0.037, 95% CI 0.36-0.97). There were no risk factors associated with the persistence of dysosmia (**Appendix 6a, 6b**). The risk of developing hair loss was associated with female sex (OR 3.00, *p*<0.001, 95% CI 1.77-5.09) (**Appendix 7**). Persistence of any symptoms was associated with female sex (coefficient 38.0, *p*=0.003, 95% CI 13.3-62.8), being diagnosed with pneumonia (coefficient 19.0, *p*=0.019, 95% CI 3.11-35.0), and severe severity compared to mild severity (coefficient 157, *p*<0.001, 95% CI 84.4-229). (**Figure 4**).

**Figure 4.**
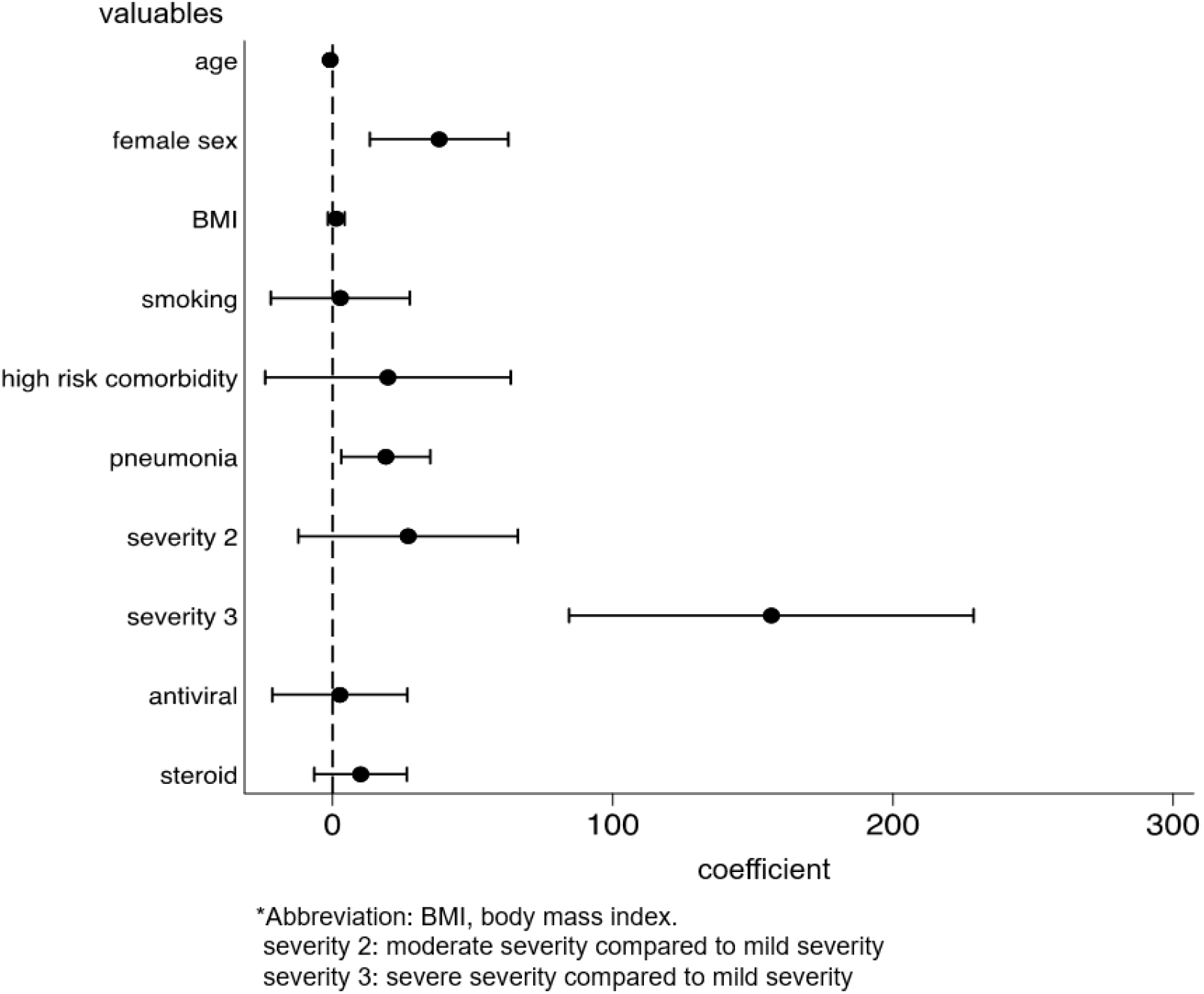
Risk factors for persistence of any symptoms associated with COVID-19.

## Discussion

This cross-sectional questionnaire survey is one of the few studies to identify the risk factors for both development and persistence of COVID-19. We also examined the duration of long COVID in more detail by surveying participants who had recovered from COVID-19 for a median period of 248.5 days.

One of the most important findings in this study was that female sex was a risk factor for developing multiple symptoms such as fatigue, dysosmia, dysgeusia, and hair loss after acute COVID-19, and were at risk for persistence of dysgeusia and any symptoms associated with COVID-19. Female sex has been recognized as a risk factor for multiple long COVID symptoms in previous studies [2][13][14]. In a cohort study in Wuhan, China, female sex was associated with the development of lung diffusion impairment, anxiety or depression, and fatigue or muscle weakness [2]. In addition, women are more prone to having multiple symptoms and are more likely to experience fatigue, sleep disorders, and mental symptoms [14]. Based on the previous literature [15], we believe that the effect of reporting bias that women are more likely to report symptoms than men is small.

Chronic fatigue is the most frequently reported symptom following recovery from acute COVID-19 [3] [16] [17]; however, its cause, pathogenesis, and reason why women are more affected than men is unclear [18]. A cross-sectional analytical study found no association between pro-inflammatory markers and chronic fatigue in COVID-19 patients [17]. Multiple factors are likely to play a role in the development of post-COVID-19 fatigue [18]. For example, a narrative review explains that congestion of the glymphatic system and the subsequent toxic build-up within the central nervous system, caused by an increased resistance to cerebrospinal fluid drainage through the cribriform plate as a result of olfactory neuron damage, may contribute to post-COVID-19 fatigue [19]. Moreover, negative psychological and social factors associated with the COVID-19 pandemic have also been linked to chronic fatigue [20] [21]. As women are more likely to be affected by anxiety, depression, and distress [22] [23], this could explain why women are more affected by chronic fatigue than men. In addition, direct severe acute respiratory syndrome coronavirus 2 (SARS-CoV-2) infection of skeletal muscle, resulting in muscle damage and weakness, may contribute to fatigue [24]. Further research is needed to explore and clarify the pathophysiology of chronic fatigue in COVID-19 patients [17].

Another important finding in this study was that younger age and lower BMI were risk factors for dysgeusia and dysosmia. Our telephone interviews conducted in Japan also revealed that younger participants, especially those in their twenties, had a higher incidence of dysosmia and dysgeusia [25]. A possible mechanism of gustatory dysfunction in COVID-19 concerns the functional link between taste and smell, whereby gustatory perception is reduced because of antecedent olfactory sensory dysfunction [26]. Long-lasting dysosmia and dysgeusia are critical issues that can affect the quality of life, even in young patients. Further research is needed to explore the pathophysiology and eventually identify treatment options.

Our study revealed that majority of participants with alopecia had diffuse hair loss. Previous studies stated that majority of patients with alopecia after COVID-19 recovery were due to telogen effluvium [27]. As telogen effluvium is a noninflammatory alopecia that causes diffuse hair loss, our findings were consistent with the previous study. On the other hand, a few participants in our study had patchy hair loss. Some case reports demonstrated that alopecia areata is a possible mechanism of patchy hair loss after COVID-19 recovery [28][29].

Lastly, the third important finding in this study was that there was little effect of antiviral medication or steroids on long COVID symptoms, except for dysosmia. The efficacy of rehabilitation has been suggested for the treatment of long COVID symptoms, but nothing has been established as effective for treatment, and the efficacy of vitamin C has also been denied [30]. Therefore, to prevent long COVID, it is important not to be infected with SARS-CoV-2.

Further research on the treatment of long COVID symptoms is also needed. On the other hand, vaccination compared to no vaccination was associated with reduced odds of long-duration (≥28 days) symptoms related to COVID-19 [31]. This implies that two doses of vaccination may shorten the duration of a long COVID. Thus, vaccination would be effective in preventing long COVID as well as, protecting oneself from the virus, and decreasing mortality.

Our study also clarified that the number of participants with at least one symptom after six months and 12 months after symptom onset or diagnosis of COVID-19 were 120 (26.3%) and 40 (8.8%), respectively. This epidemiological data revealed that most patients with long COVID recovered over one year. With the risk factors associated with persistence of fatigue, dysosmia, dysgeusia, or hair loss, patients with these symptoms can predict the duration of each symptom, which might reduce the anxiety of not knowing how long each symptom would continue. Longitudinal follow-up surveys are needed to better understand the natural history of long COVID.

Our study has several limitations. First, this study relied on patient self-reports and, therefore, might have been subject to recall bias. Second, some patients reported on-going/chronic and late-onset symptoms at the time of the questionnaire survey. In these cases, the actual duration of the symptoms was unclear, and it is likely that this study underestimated the duration of these symptoms. Long-term observation is needed to better understand the duration of a long COVID. Third, although a previous study indicated that severity of illness was a risk factor for developing fatigue [2], being severe was not a risk factor for developing fatigue in this study. This is partly because most of the participants in our study were mild cases, and the number of severe cases was markedly small. Further studies are needed to clarify whether the severity of illness is a risk factor for the development and persistence of long COVID.

In conclusion, our cross-sectional questionnaire survey evaluated patients after recovery from COVID-19, most of whom had mild disease, for the development and persistence of long COVID. Women were at risk for development of fatigue, dysosmia, dysgeusia, and hair loss, and were at risk for persistence of dysgeusia and any symptoms associated with COVID-19. Younger age and low BMI were found to be at risk of developing dysosmia and dysgeusia. In addition, about a quarter of the participants had at least one prolonged symptom for more than six months, indicating that many COVID-19 patients suffer from long-term residual symptoms, even in mild cases.

## Data Availability

The authors confirm that the data supporting the findings of this study are available within the article and its supplementary materials.

## Acknowledgment

We thank all the people who participated in our study, “Collection and antibody measurement of Convalescent plasma foreseeing the use for COVID-19 treatment”.

