## Appendix 1 for "Risk factors associated with development and persistence of long COVID"

**Appendix 1. Questionnaire**

If you consent to participate in this survey, please put a check mark in the □ below and write the date on which you responded to the survey.

**□ I hereby consent to participate in this study.**

**Response date (using the format Month/Day/Year): _____/_____/2021**

1) The following questions are about you. For multiple-choice questions, please circle the answer that applies to you. For questions that include parentheses (      ), please write your answer inside the parentheses.

Age （ ） years

Sex

1. Male
2. Female

Height （ ） cm

Weight （ ） kg

History of smoking

3. Yes
4. No

History of alcohol consumption

1. Yes
2. No

Medical history (past illnesses)

1. Hypertension
2. Diabetes
3. Dyslipidemia
4. Bronchial asthma
5. Chronic obstructive pulmonary disease (COPD)
6. Myocardial infarction

7. Melanoma
8. Connective tissue/rheumatic disease
9. Immune deficiency disease
10. Chronic kidney disease
11. Other（　　　　　）

Obstetric history (women)

1. I had a pregnancy before I was infected with coronavirus.
2. I was pregnant when I was infected with coronavirus.
3. I have never been pregnant.

2) The following questions are about the course of your coronavirus infection. Please circle the answers that apply to you.

| | | | |
|---|---|---|---|
| I was diagnosed with pneumonia | 1. Yes | 2. No | 3. I don't know |
| I was administered oxygen once or more | 1. Yes | 2. No | 3. I don't know |
| I was put on a ventilator once or more | 1. Yes | 2. No | 3. I don't know |
| I was treated with ECMO once or more | 1. Yes | 2. No | 3. I don't know |
| I was prescribed antivirals[1] | 1. Yes | 2. No | 3. I don't know |
| I was prescribed steroids[2] | 1. Yes | 2. No | 3. I don't know |

Notes: 1) Antivirals are drugs used to COVID-19, such as remdesivir (Veklury), favipiravir (Avigan), ivermectin (Stromectol), lopinavir/ritonavir (Kaletra), hydroxychloroquine (Plaquenil), and azithromycin (Zithromax).
2) Steroids refer to drugs that inhibit inflammation, such as dexamethasone (Dexart), prednisolone (Predonin), and methylprednisolone (Solu-Medrol).

3) The following questions are about the aftereffects of COVID-19. Please note whether you experienced the following symptoms. If yes, please write the duration of that symptom in days. If you are unsure, please write the approximate duration.

Example: If you had a symptom for 20 days starting on day 30 after coronavirus infection, please respond as follows:

1. Yes 2. No

1. If yes, A. Since infection B. Since day（ 30 ） after infection

a. For （20 ） days b. Ongoing

**Fever**

1. Yes 2. No

1. If yes, A. Since infection B. Since day （ ）after infection

a. For（ ）days b. Ongoing

**Fatigue (Sluggishness)**

1. Yes 2. No

1. If yes, A. Since infection B. Since day （ ）after infection

a. For（ ）days b. Ongoing

**Shortness of breath (Difficulty breathing)**

1. Yes 2. No

1. If yes, A. Since infection B. Since day （ ）after infection

a. For（ ）days b. Ongoing

**Joint pain**

1. Yes 2. No

1. If yes, A. Since infection B. Since day （ ） after infection

a. For （ ） days b. Ongoing

**Myalgia (Muscle pain)**

1. Yes 2. No

1. If yes, A. Since infection B. Since day （ ） after infection

a. For （ ） days b. Ongoing

**Chest pain**

1. Yes 2. No

1. If yes, A. Since infection B. Since day （ ） after infection

a. For （ ） days b. Ongoing

**Cough**

1. Yes 2. No

1. If yes, A. Since infection B. Since day （ ） after infection

a. For （ ） days b. Ongoing

**Abdominal pain (Stomach pain)**

1. Yes 2. No

1. If yes, A. Since infection B. Since day （ ） after infection

a. For（　　　　）days　　　　b. Ongoing

**Dysosmia (loss of smell)**

1. Yes　　　　2. No

1. If yes, A. Since infection　　　　B. Since day （　　　　）after infection

a. For（　　　　）days　　　　b. Ongoing

**Dysgeusia (loss of taste)**

1. Yes　　　　2. No

1. If yes, A. Since infection　　　　B. Since day （　　　　）after infection

a. For（　　　　）days　　　　b. Ongoing

**Nasal discharge**

1. Yes　　　　2. No

1. If yes, A. Since infection　　　　B. Since day （　　　　）after infection

a. For（　　　　）days　　　　b. Ongoing

**Bloodshot eyes**

1. Yes　　　　2. No

1. If yes, A. Since infection　　　　B. Since day （　　　　）after infection

a. For（　　　　）days　　　　b. Ongoing

**Headache**

1. Yes 2. No

1. If yes, A. Since infection B. Since day （ ） after infection

a. For（ ） days b. Ongoing

**Phlegm**

1. Yes 2. No

1. If yes, A. Since infection B. Since day （ ） after infection

a. For（ ） days b. Ongoing

**Sore throat**

1. Yes 2. No

1. If yes, A. Since infection B. Since day （ ） after infection

a. For（ ） days b. Ongoing

**Diarrhea**

1. Yes 2. No

1. If yes, A. Since infection B. Since day （ ） after infection

a. For（ ） days b. Ongoing

**Nausea/vomiting**

1. Yes 2. No

1. If yes, A. Since infection B. Since day （ ） after infection

a. For（ ） days b. Ongoing

**Loss of appetite**

1. Yes 2. No

1. If yes, A. Since infection B. Since day （ ） after infection

a. For（ ） days b. Ongoing

**Hair loss**

1. Yes 2. No

1. If yes, A. Since infection B. Since day （ ） after infection

a. For（ ） days b. Ongoing

i. In patches ii. Diffuse hair loss iii. Don’t know

**Depression**

1. Yes 2. No

1. If yes, A. Since infection B. Since day （ ） after infection

a. For（ ） days b. Ongoing

**Loss of concentration**

1. Yes 2. No

1. If yes, A. Since infection B. Since day （ ） after

infection

a. For（　　　　）days　　　　b. Ongoing

**Memory disturbance (forgetfulness)**

1. Yes　　　　2. No

1. If yes, A. Since infection　　　　B. Since day （　　　　）after infection

a. For（　　　　）days　　　　b. Ongoing

**Appendix 2. The number of the participants with COVID-19 symptoms and their persistence**

| symptoms | Number (%[a]) | Lasting more than 4 weeks (%[a]) | Lasting more than 12 weeks (%[a]) | Late onset (%[b]) |
|---|---|---|---|---|
| fever | 293 (64.1) | 12 (2.6) | 1 (0.2) | 2 (0.7) |
| fatigue | 292 (64.0) | 93 (20.4) | 47 (10.3) | 8 (2.7) |
| dysosmia | 219 (47.9) | 104 (22.8) | 62 (13.6) | 4 (1.8) |
| cough | 214 (46.8) | 68 (14.9) | 21 (4.6) | 4 (1.9) |
| dysgeusia | 185 (40.6) | 69 (15.1) | 30 (6.6) | 2 (1.1) |
| headache | 173 (37.9) | 32 (7.0) | 18 (3.9) | 3 (1.7) |
| LoA | 171 (37.6) | 24 (5.3) | 9 (2.0) | 0 (0) |
| SoB | 163 (35.7) | 55 (12.0) | 25 (5.5) | 4 (2.5) |
| joint pain | 153 (33.5) | 20 (4.4) | 15 (3.3) | 5 (3.3) |
| sore throat | 139 (30.5) | 12 (2.6) | 5 (1.1) | 1 (0.7) |
| LoC | 137 (30.0) | 95 (20.8) | 70 (15.3) | 10 (7.3) |
| depression | 125 (27.5) | 79 (17.3) | 55 (12.0) | 16 (12.8) |
| myalgia | 113 (24.7) | 16 (3.5) | 11 (2.4) | 3 (2.7) |
| diarrhea | 111 (24.3) | 6 (1.3) | 4 (0.9) | 2 (1.8) |

| | | | | |
|---|---|---|---|---|
| sputum | 109 (23.9) | 25 (5.5) | 13 (2.8) | 2 (1.8) |
| hair loss | 103 (22.7) | 73 (16.0) | 29 (6.3) | 58 (56.3) |
| MD | 102 (22.5) | 88 (19.3) | 78 (17.1) | 17 (16.7) |
| runny nose | 96 (21.1) | 14 (3.1) | 9 (2.0) | 0 (0) |
| chest pain | 92 (20.1) | 32 (7.0) | 18 (3.9) | 7 (7.6) |
| abdominal pain | 47 (10.3) | 3 (0.7) | 1 (0.2) | 0 (0) |
| nausea | 36 (7.9) | 4 (0.9) | 3 (0.7) | 0 (0) |
| conjunctivitis | 32 (7.0) | 4 (0.9) | 1 (0.2) | 0 (0) |

Abbreviations: LoA, loss of appetite; SoB, shortness of breath; LoC, loss of concentration; MD, memory disturbance.

[a]Calculated by dividing the number of patients by the total number of participants (n =457).

[b]Calculated by dividing the number of patients by the number of participants who experienced the symptoms (N=457).

### Appendix 3. Acute symptoms associated with COVID-19

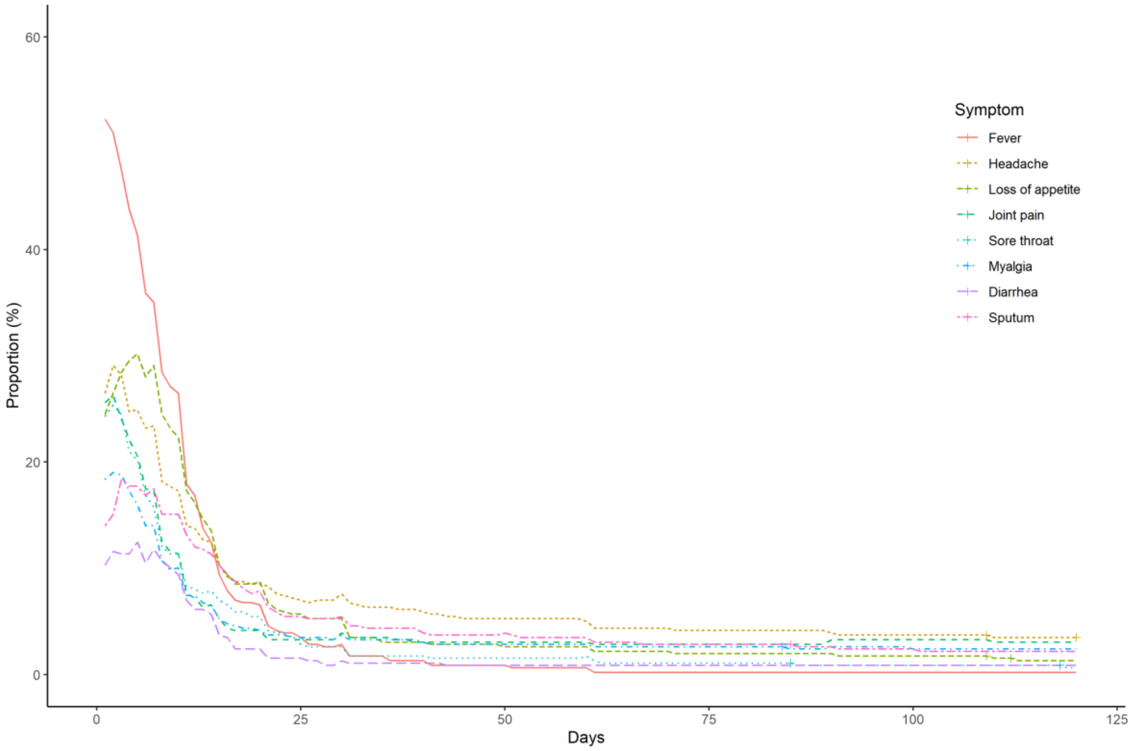

**Appendix 4a. Risk factors for development of fatigue in patients with COVID-19**

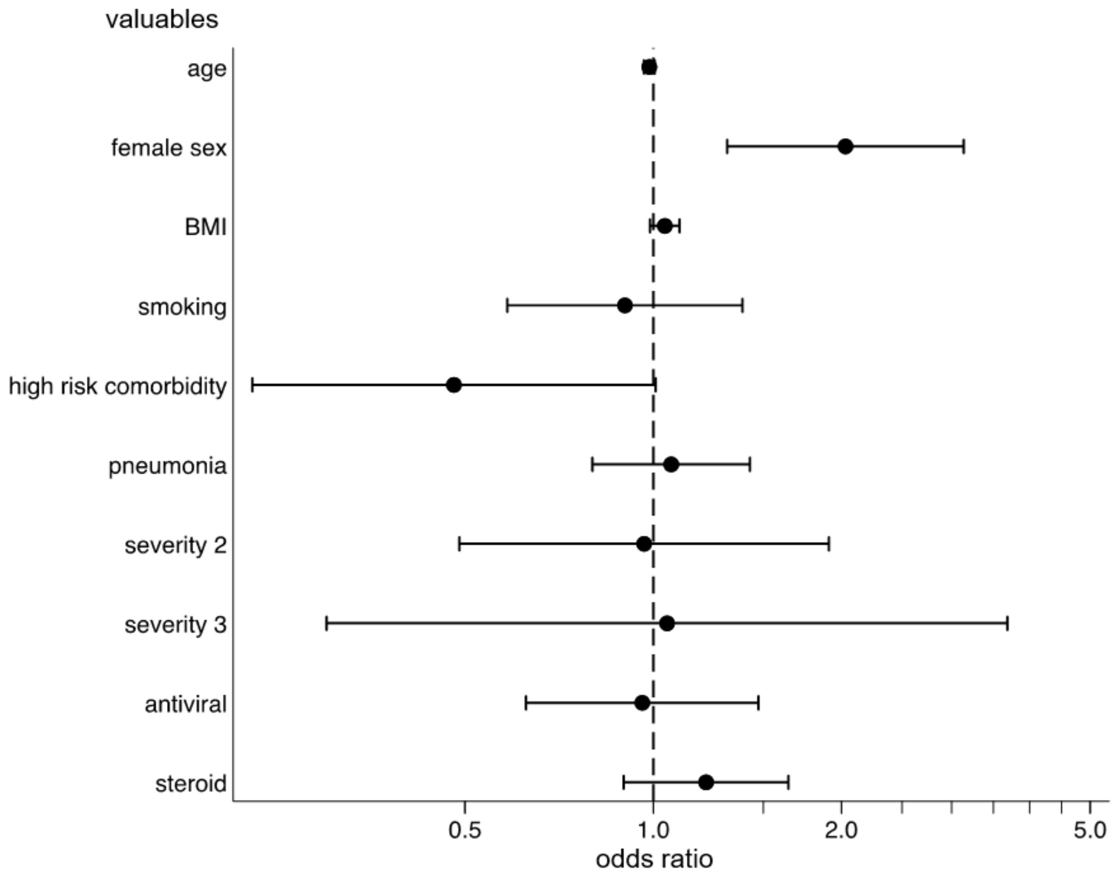


*Abbreviation: BMI, body mass index.
severity 2: moderate severity compared to mild severity
severity 3: severe severity compared to mild severity

**Appendix 4b. Risk factors for persistence of fatigue among patients with fatigue**

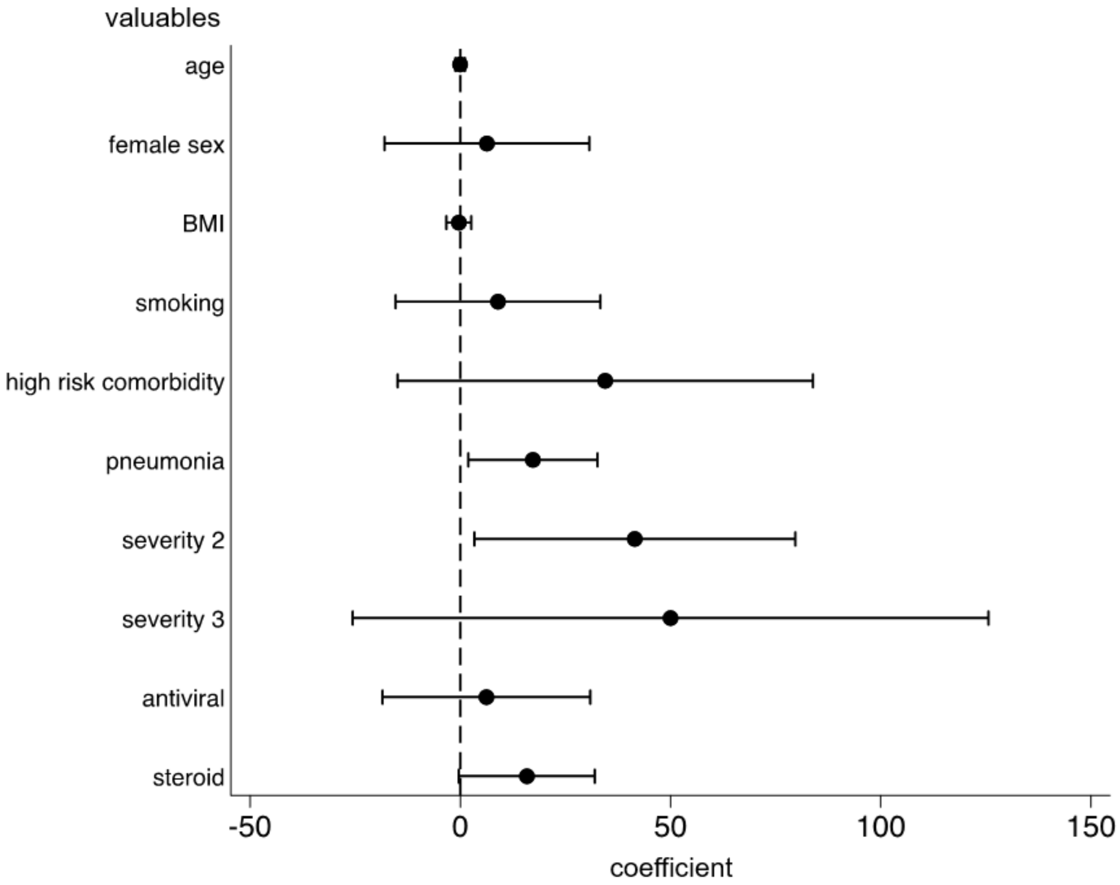


*Abbreviation: BMI, body mass index.
severity 2: moderate severity compared to mild severity
severity 3: severe severity compared to mild severity

**Appendix 5a. Risk factors for development of dysgeusia in patients with COVID-19**

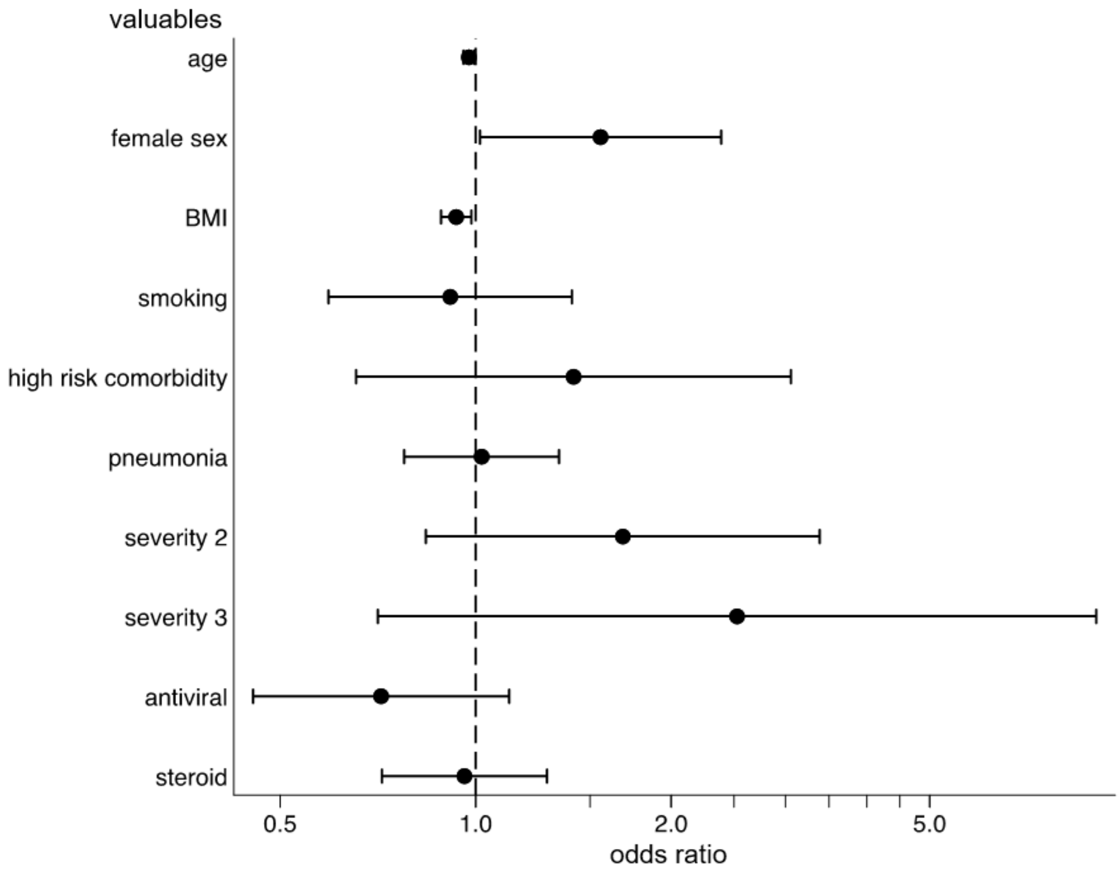


*Abbreviation: BMI, body mass index.
severity 2: moderate severity compared to mild severity
severity 3: severe severity compared to mild severity

**Appendix 5b. Risk factors for persistence of dysgeusia among patients with dysgeusia**

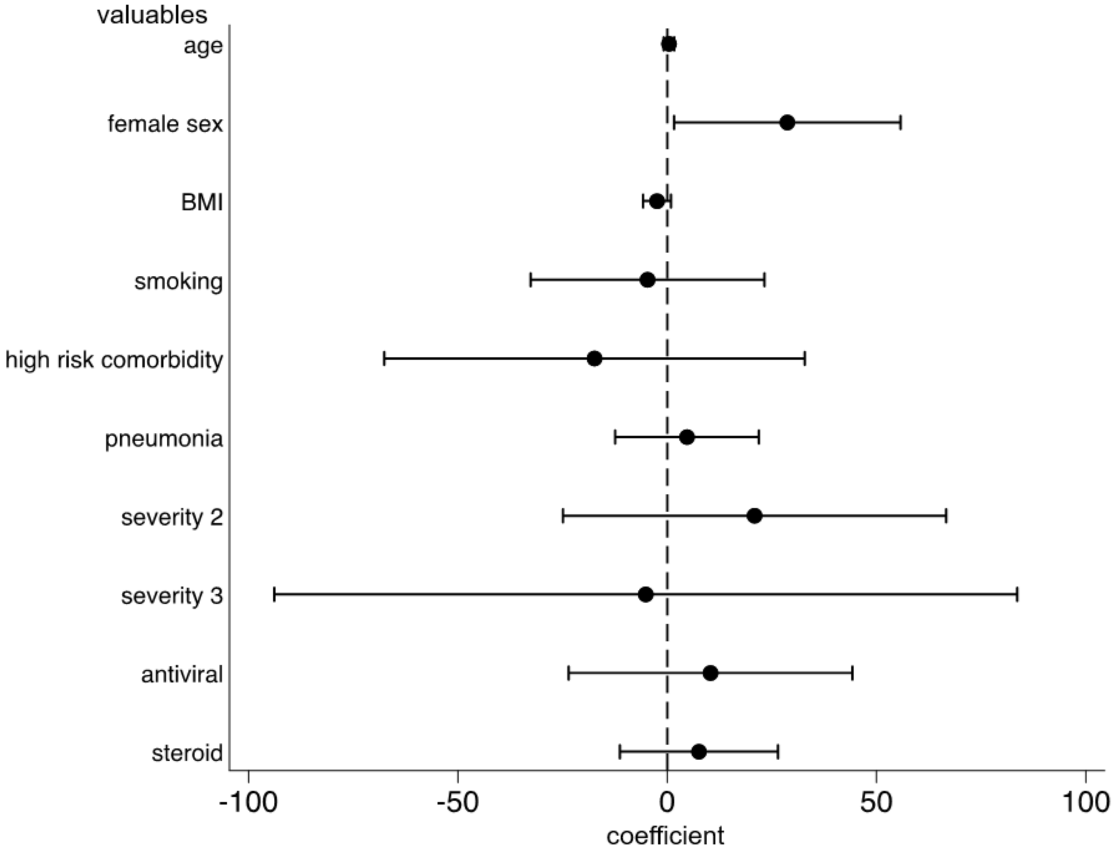


*Abbreviation: BMI, body mass index.
severity 2: moderate severity compared to mild severity
severity 3: severe severity compared to mild severity

**Appendix 6a. Risk factors for development of dysosmia in patients with COVID-19**

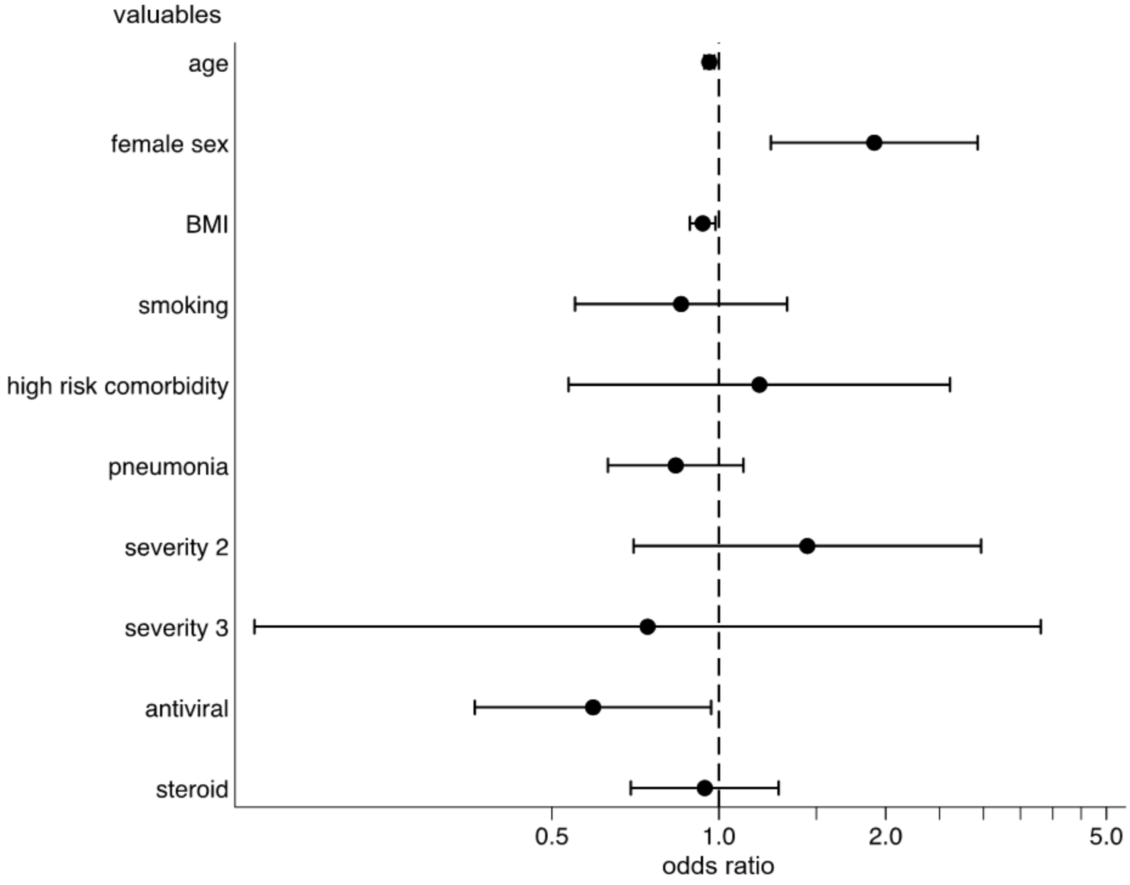


*Abbreviation: BMI, body mass index.
severity 2: moderate severity compared to mild severity
severity 3: severe severity compared to mild severity

**Appendix 6b. Risk factors for persistence of dysosmia among patients with dysosmia**

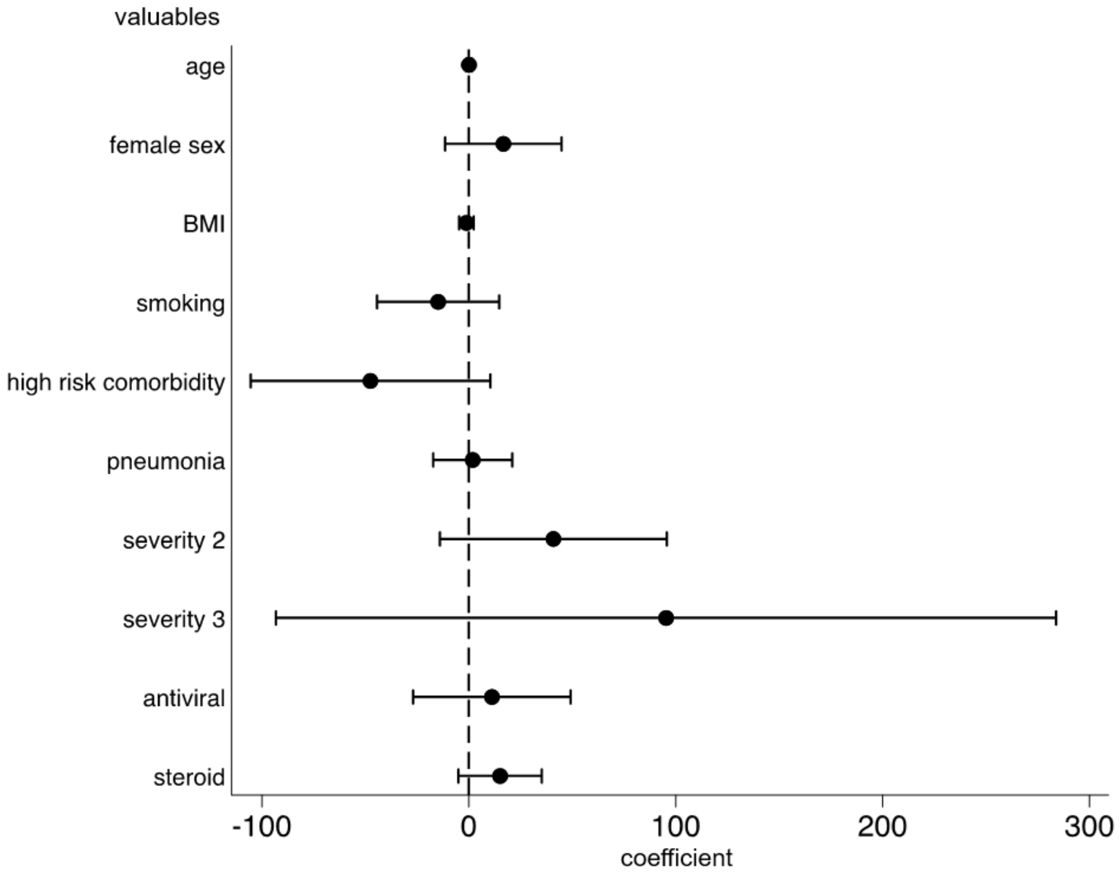


*Abbreviation: BMI, body mass index.
severity 2: moderate severity compared to mild severity
severity 3: severe severity compared to mild severity

**Appendix 7. Risk factors for development of hair loss in patients with COVID-19**

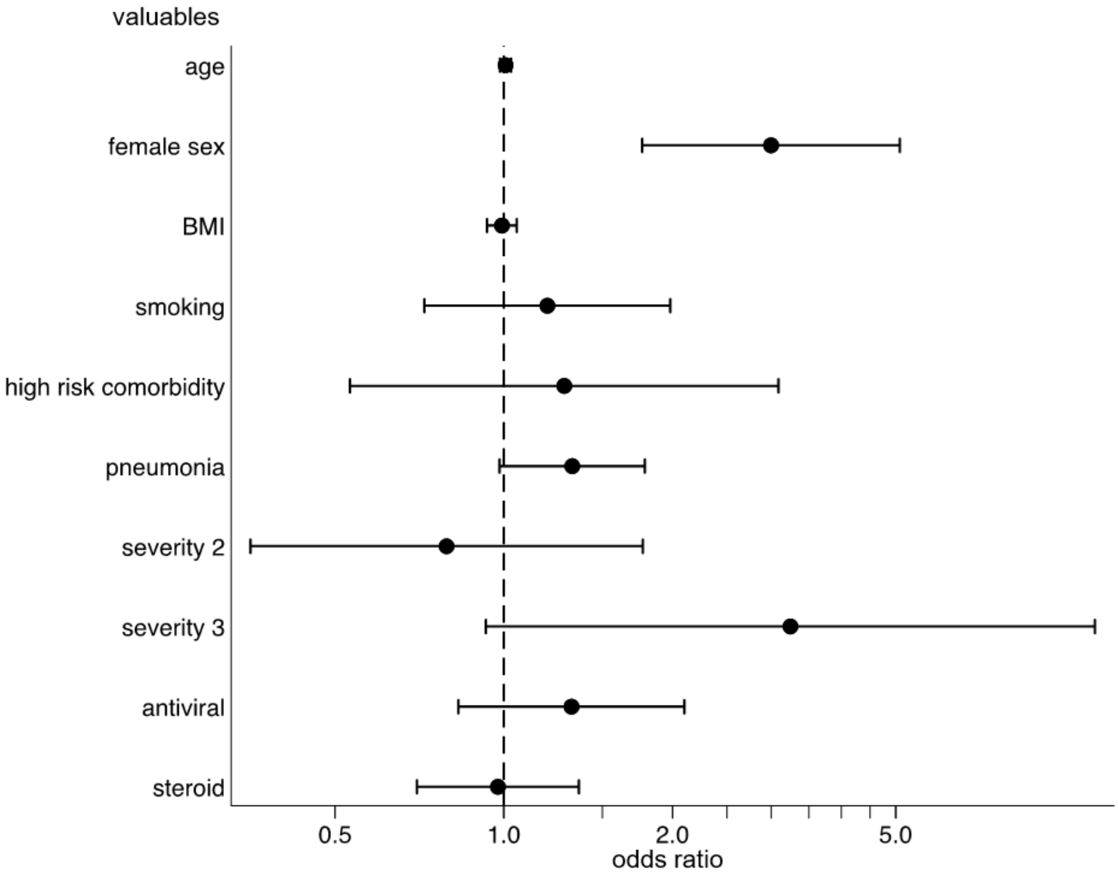


*Abbreviation: BMI, body mass index.
severity 2: moderate severity compared to mild severity
severity 3: severe severity compared to mild severity
